# Choroid Plexus Enlargement in Secondary Progressive MS: phenotype comparison

**DOI:** 10.1101/2025.03.31.25324925

**Authors:** Samuel Klistorner, Michael H Barnett, Chenyu Wang, Anneke Van der Walt, Helmut Butzkueven, Zhaoyuan Gong, Mustapha Bouhrara, John Parratt, Con Yiannikas, Alexander Klistorner

## Abstract

**Background:** The choroid plexus (CP) is increasingly recognised as a contributor to chronic inflammation in multiple sclerosis (MS). While CP enlargement is reported in early MS, its role in secondary progressive MS (SPMS) is poorly understood.

**Objective:** We aimed to quantify CP volume in SPMS and compare it to relapsing-remitting MS (RRMS) and clinically isolated syndrome (CIS), and to assess associations with disease severity and progression.

**Methods:** CP volumes were manually segmented and normalised to intracranial volume. Age correction was applied using a healthy control cohort. Cross-sectional and longitudinal analyses evaluated relationships with ventricular volume, lesion burden, and brain atrophy.

**Results:** CP volume increased significantly across MS phenotypes: SPMS patients showed 26% higher CP volume than CIS (p=0.010) and 17% higher than RRMS (p=0.034). CP enlargement in SPMS was independent of ventricular size, indicating distinct underlying mechanisms. While lesion burden was the primary determinant of brain atrophy in SPMS, longitudinal data revealed significant associations between CP volume, chronic lesion expansion (r^2^=0.31), and brain volume loss (r^2^=0.52).

**Conclusion:** CP enlargement is a progressive feature of MS, not driven by ventricular expansion. In SPMS, it may reflect ongoing inflammation contributing to tissue damage, supporting its role as a biomarker.

## Introduction

The choroid plexus (CP), a key component of the blood-cerebrospinal fluid barrier, plays an integral role in immune regulation within the central nervous system (CNS) [1][2][3][4][5][6][7][8][9][10][11][12][13][14][15][16][17][18]; and has recently emerged as a focus of interest in multiple sclerosis (MS) research. Increases in CP volume have been reported in patients with relapsing-remitting MS (RRMS) and even in those with clinically or radiologically isolated syndrome (CIS)[19][12], suggesting its involvement in both early and established stages of MS. This volumetric enlargement has been linked to heightened chronic inflammation and immune activation, with transient enlargement observed during acute inflammatory relapses. Despite growing evidence of CP involvement in MS pathophysiology, volumetric changes in the CP and associated clinical implications remain unexplored in secondary progressive multiple sclerosis (SPMS).

SPMS represents a later stage of MS characterised by reduced inflammatory relapses and a shift towards progressive neurodegeneration, making it critical to investigate whether CP volume dynamics differ from those observed earlier in the disease. Understanding the role of the CP in SPMS could provide novel insights into its contribution to disease progression and severity.

In this study, we aim to quantify CP volume in SPMS patients and compare it to RRMS and CIS cohorts. Additionally, we seek to explore the relationship between CP volume and clinical and radiological markers of disease severity. Finally, we investigate whether CP enlargement is secondary to ventricular expansion due to its mechanical attachment to the ventricles, as recently suggested.[5]

## Methods

### Subjects

We analysed data from three distinct MS cohorts and a healthy control group. The primary cohort comprised SPMS patients enrolled in a longitudinal study (MS Australia grant, 21-4-010). The diagnosis of SPMS was determined by the treating neurologist [20][21]

For comparative analyses, patients with established relapsing-remitting multiple sclerosis (RRMS), diagnosed according to the revised McDonald 2010 criteria [22] were recruited from the Mechanisms of Axonal Degeneration in MS study (MADMS, National MS Society grant, G-1508-05946), Additionally, CIS patients were included from a prospective optic neuritis (ON) study conducted at the Royal Melbourne Hospital between 2008 and 2010. All CIS patients presented with acute unilateral ON, diagnosed using standardised clinical criteria identical to those employed in the Optic Neuritis Treatment Trial. [23] Clinical follow-up data were available at 3 years and 10 years post-presentation. Only patients who progressed to clinically definite multiple sclerosis within the 10-year follow-up period were included in the current study.

The control cohort comprising neurologically healthy subjects spanning a broad age range, was drawn from two studies, the Baltimore Longitudinal Study of Aging (BLSA) and the Genetic and Epigenetic Signatures of Translational Aging Laboratory Testing (GESTALT) study [24] This cohort enabled the development of age-correction algorithms for choroid plexus and ventricular measurements in MS patients and facilitated the investigation of the normative relationship between these structures. This analysis was particularly relevant for the SPMS cohort, where advanced age was frequently observed.

### CP measurement

Choroid plexus (CP) within the lateral ventricles were manually segmented in MS patients using post-gadolinium (GAD) T1-weighted images on JIM 9 software (Xinapse Systems, Essex, UK). All segmentations were performed by a trained analyst (AK). CP volumes were normalised as a percentage of total intracranial volume (TIV) and scaled by a factor of 1000 to enhance interpretability. [25] For the control cohort, CP segmentation was performed on T1 images due to the absence of GAD-enhanced imaging and was only used for calculating age-related changes. [24]

### Brain volumetrics

Brain volumetric analyses were performed on T1-weighted images utilising AssemblyNet, an artificial intelligence (AI)-based brain segmentation tool [26]. All volumetric measurements were normalised and expressed as a percentage of total intracranial volume (TIV), enabling meaningful cross-sectional comparisons across participants by controlling for natural variations in head size.

### Age adjustments

Age-related effects were accounted for using normative data from the healthy control cohort. For CP and ventricular volumes, age-volume relationships were modelled using quadratic functions derived from control data [24]. Brain volume age adjustment was implemented following the linear correction method described by Whitwell et al. [26] for subjects over 35 years. Detailed age adjustment methodology and formulae are provided in Supplementary Materials. This standardisation approach effectively accounted for age-related variability whilst preserving sensitivity to disease-specific alterations. All subsequent analyses utilised these age-adjusted values to ensure robust characterisation of disease-related changes.

### Lesion Masking

Lesion masking was performed utilising iQ-Solutions™-MS Report (Sydney Neuroimaging Analysis Centre, Sydney, Australia) [27], a state-of-the-art AI-based analytical suite using T1-weights and FLAIR sequences as inputs.

### Statistics

Between-group comparisons of demographic and clinical characteristics were performed using appropriate statistical tests based on data distribution. Analysis of variance (ANOVA) was employed to assess age differences between clinical phenotypes, with subsequent post-hoc analyses for paired group comparisons. Gender distribution across groups was evaluated using the chi-square test.

For volumetric analyses, one-way ANOVA was employed to assess between-group differences in age-adjusted choroid plexus volumes, age-adjusted ventricular volumes, age-adjusted brain volumes, and total lesion volumes. Post-hoc analyses utilising Tukey’s Honest Significant Difference (HSD) test were conducted for all paired group comparisons.

Non-parametric Kruskal-Wallis H-test was utilised to analyse between-group differences in EDSS scores, with subsequent post-hoc analyses for paired comparisons.

Relationships between volumetric measures were assessed using Pearson’s correlation coefficient, with partial correlations employed to control for age effects. The influence of age on the relationship between choroid plexus and ventricular volumes was further investigated utilising mediation analysis with the Bootstrap Resampling Method.

Multiple linear regression analyses were performed to evaluate predictors of brain volume. A comprehensive model incorporating choroid plexus volume, total lesion volume, age, and gender was developed for the entire cohort. Subsequently, phenotype-specific models were constructed to examine predictor patterns within individual clinical groups. Standardised beta coefficients (β) were calculated to enable direct comparison of predictor contributions across different measurement scales.

Longitudinal analyses were conducted on a subset of SPMS patients with 5-year follow-up data. Linear regression was employed to examine associations between choroid plexus volume and both annual rates of chronic lesion expansion and brain volume reduction.

Annual rates were calculated as percentage changes from baseline measurements.

All statistical analyses were performed using Python statistical packages. Statistical significance was set at p<0.05, with appropriate corrections applied for multiple comparisons.

## Results

### Group comparison

Demographic and clinical characteristics of the patient cohorts are summarised in Table 1. Analysis of baseline data revealed several major differences between clinical phenotypes. SPMS patients were significantly older and demonstrated markedly higher lesion volumes compared to both CIS and RRMS groups (p<0.0001 and p<0.001, respectively). While gender distribution showed a trend towards higher male representation in SPMS, this did not reach statistical significance. Normalised Brain Volume (NBV) differed significantly across all phenotypes (p<0.001 for all paired comparisons), while ventricular enlargement was notably more pronounced in SPMS compared to both CIS and RRMS cohorts (p=0.002 and p=0.006, respectively). EDSS scores were significantly elevated in the SPMS cohort compared to both CIS and RRMS groups (p<0.001).

Analysis of CP volumes revealed significant differences across clinical phenotypes (univariate ANOVA, p=0.008). Post-hoc analyses demonstrated significantly enlarged CP volumes in the SPMS cohort compared to CIS (p=0.010) and RRMS groups (p=0.034), but no difference between CIS and RRMS (p=0.587)

### Relationship between CP volume and ventricular size

A strong link between CP and lateral ventricular volume was observed in normal controls (r^2^ = 0.65, p<0.001). The correlation, however, significantly decreased after adjusting for age (partial correlation: r^2^ = 0.30, p<0.001) (see Suppl Fig1).

This is also supported by mediation analysis (using Bootstrap Resampling Method), which demonstrated a significant effect of both direct (i.e. ventricular size, 0.021, p<0.001) and indirect (i.e. age, 0.009, p<0.001) factors on plexus volume in normal controls. This analysis shows that age explains a considerable part of the relationship between the CP and ventricles. (0.0087 / 0.0299 or 29% of the total effect).

Analysis of the relationship between CP and lateral ventricular volumes performed using age-adjusted values (as described in Method) revealed distinct patterns across clinical phenotypes. In the SPMS cohort, correlation analysis between CP and ventricular volumes showed no significant association (r^2^=0.04, p=0.239). Notably, some patients exhibited disproportionately enlarged ventricles with modest CP volumes (Fig.2 c, blue circle, or examples in Fig3. a, b), whilst others demonstrated moderately sized ventricles with markedly enlarged CP volumes (Fig.2c, green circle or examples in Fig.3c, d).

Conversely, the RRMS cohort displayed a moderate but significant positive correlation between age-adjusted CP and ventricular volumes (r^2^=0.29, p<0.001), aligning with the strength of the relationship between the two observed in the normal controls (Fig 2b).

The CIS cohort demonstrated no significant association between the two structures and was characterised by variable choroid plexus volumes accompanying predominantly smaller ventricular sizes (r^2^=0.00 p=0.91) (Fig.2a).

### Correlation of CP with Clinical and Radiological measures of disease severity

Analyses of relationships between choroid plexus volume and established disease metrics revealed distinct patterns across clinical phenotypes, as shown in Figure 4. In the SPMS cohort, choroid plexus volume demonstrated no significant associations with disability measures (EDSS: r^2^=0.03, p=0.25), total brain T2 lesion burden (r^2^=0.00, p=0.869), or NBV (r^2^=0.01, p=0.576).

Similarly, the CIS cohort exhibited no significant correlations between choroid plexus volume and either lesion burden (r^2^=0.01 p=0.622) or NBV (r^2^=0.06, p=0.173).

In contrast, volumetric analyses in the RRMS cohort revealed significant associations between choroid plexus volume and both total brain T2 lesion burden (r^2^=0.24, p<0.001) and NBV (r^2^=0.20, p<0.001).

### Multivariate Analysis of Brain Volume Predictors

The model of brain volume prediction incorporating choroid plexus volume, total lesion volume, age, and gender, across the entire dataset (including CIS, RRMS, and SPMS patients), demonstrated high predictive power (R^2^=0.78). Plexus volume was negatively associated with lesion volume (β=−2.19, p<0.001), brain volume (β=−0.68, p=0.004) and age (β=−1.62, p<0.001) and demonstrated a weak but significant association with sex (β=−0.85, p=0.047).

Multiple linear regression analyses examining predictors of brain atrophy in individual cohorts revealed varying model performance across clinical phenotypes. The model demonstrated strong predictive capability for SPMS (R^2^=0.70, p<0.001) and more moderate power for RRMS cohorts (R^2^=0.61, p<0.001), whilst yielding minimal explanatory power in CIS patients (R^2^=0.27, p=0.16).

Analysis of standardised coefficients (β) demonstrated the relative contributions of individual predictors (Table.2). The RRMS cohort analysis revealed significant contributions from both total lesion volume (β=−0.443, p<0.001) and choroid plexus volume (β=−0.218, p=0.02), with lesion volume demonstrating a 2-fold stronger predictive value. Age similarly emerged as a significant predictor of reduced brain volume (β=−0.371, p<0.001).

In the SPMS cohort, lesion burden emerged as the dominant predictor of NBV (β=−0.651, p<0.001), with its contribution exceeding that of choroid plexus volume by a factor of 5. An additional significant predictor was male sex (β=−0.285, p=0.008). Notably, choroid plexus volume demonstrated no significant predictive value (β=−0.129, p=0.27).

### Longitudinal Analysis of CP Volume and Disease Progression in SPMS

Longitudinal MRI data spanning a 5-year period were available for a subset of SPMS patients (n=13/40), acquired using consistent scanner and protocol parameters. Linear regression analyses revealed significant associations between choroid plexus volume and both the annual rate of chronic lesion expansion (r^2^=0.31, p=0.04; mean expansion rate 18.1±12.3% per year) and annual brain volume reduction (r^2^=0.52, p=0.006), as shown in Supplementary figure 2.

## Discussion

The CP, a key interface between the immune system and brain, is increasingly recognised as a contributor to neuroinflammation and MS progression. Its roles in cerebrospinal fluid production and immune cell trafficking make it a potential biomarker of disease activity and severity.

In this study, we assessed CP volume in SPMS patients, comparing it with other MS phenotypes and examining its relationship with ventricular size and disease severity. To our knowledge, this is the first study to specifically evaluate CP volume in SPMS, offering new insights into its role in advanced disease stages.

Our analysis revealed several novel findings:

### 1. CP Volume continues to increase across disease stages

Previous studies have shown significant CP enlargement in patients with clinically isolated syndrome (CIS), radiologically isolated syndrome (RIS), and paediatric-onset MS (POMS),[19][18][5] suggesting that this process may even precede the onset of clinical symptoms.

Independent reports consistently demonstrate a 25–30% increase in CP volume in these early stages compared to healthy controls. Similar enlargement (27–35%) has also been observed in early RRMS. [16][1][28] Our current analysis shows that CP volume continues to increase as MS progresses. Specifically, age-adjusted CP volume in SPMS was significantly greater than in CIS converters and RRMS patients—by 26% and 17%, respectively.

This aligns with our previous longitudinal RRMS study, where CP volume increased steadily over time..[18] The differences observed here are consistent with the expected progression based on a previously established annual increase of 1.4% [18] and the average disease duration gap between groups (16.3 years between CIS and SPMS; 10.5 years between RRMS and SPMS, which correspond to CP volume increase of 23% and 15% respectively).

These findings highlight the progressive nature of CP enlargement across MS phenotypes and support its potential role as a biomarker of disease progression, as recently suggested.[15] CP volume may reflect ongoing immune activation and chronic inflammation within the CNS— processes increasingly recognised as key contributors to MS progression.[29]

### 2. CP enlargement in MS is not driven by ventricular expansion

The association between CP enlargement and ventricular volume, reported in several recent studies,[19][1][7][17][5] has led some researchers to propose a mechanical link between the two. A recent study suggests that as the CP is anatomically attached to the ventricular wall, progressive ventricular expansion may exert traction on the CP, causing subsequent enlargement. [5]

However, our findings, supported by earlier work,[18] challenge this hypothesis. We observed no significant correlation between CP and ventricular volumes in CIS or SPMS cohorts. Moreover, the temporal patterns of enlargement differ across disease stages.

In CIS, CP volume was significantly increased compared to healthy controls, [12] despite similar ventricular volumes. In RRMS, both CP and ventricular volumes were moderately enlarged, while in SPMS, ventricular expansion far exceeded CP growth.

Supporting this, Ricigliano et al. [19] also reported substantial CP enlargement early in the disease course, with minimal progression thereafter, while ventricular volume increased steadily across MS stages.

This dissociation points to distinct underlying mechanisms.Early CP enlargement likely reflects inflammation-related changes—such as immune activation, altered CSF turnover, or CP involvement in immune surveillance—rather than mechanical stretch from ventricle expansion. In contrast, ventricular enlargement in SPMS is primarily driven by cumulative lesion-related tissue loss. (see section 3 of Discussion).

Furthermore, In the current study, the moderate correlation between ventricular size and CP volume in the RRMS cohort closely mirrored that seen in age-adjusted healthy controls.

Since ventricular enlargement in healthy individuals is not driven by disease processes, this suggests that the observed association in RRMS likely reflects inter-individual anatomical variability rather than disease-related causality.

These findings imply that variations in CP and ventricular size in RRMS are more likely due to natural anatomical differences (similar to those seen in the normal population), rather than one structure mechanically influencing the other. This supports the view that CP enlargement is not simply a by-product of ventricular expansion, but instead reflects distinct and independent pathological or physiological processes. [29][30][15][9],

### 3. Differential Association Between Choroid Plexus Volume and Brain Atrophy Across MS Phenotypes

Our combined model—incorporating data from CIS, RRMS, and SPMS patients, and including CP volume, lesion volume, age, and gender—explained nearly 80% of the variability in brain volume. This strong result highlights several key associations.

As expected, lesion volume showed a robust negative correlation with brain volume, confirming its dominant role in driving brain atrophy. CP volume also demonstrated a significant negative association with brain volume, indicating that larger CP size is linked to greater atrophy, independent of lesion burden. Age similarly correlated with brain volume decline, reflecting well-established ageing effects. A weaker but significant association with sex suggested that males may experience more brain volume loss than females.

When analysed by phenotype, notable differences emerged. In RRMS, both lesion burden and CP enlargement significantly contributed to brain atrophy, suggesting an additive effect of focal and chronic inflammation. However, in SPMS, lesion volume was the predominant driver of tissue loss, consistent with earlier findings that high early lesion load predicts faster progression and SPMS conversion. [31][32] Indeed, the average lesion volume in SPMS patients was over three times greater than in those with RRMS, emphasising the cumulative impact of lesion burden.

Despite this, our findings also point to a continuing role for CP-mediated inflammation in SPMS. Previous work has linked CP enlargement to chronic lesion expansion and periventricular atrophy, likely driven by cytotoxic mediators diffusing from the CP via CSF, fuelling low-grade inflammation at lesion rims.[16] The present study supports this mechanism, showing a longitudinal association between CP volume, lesion expansion, and ongoing brain atrophy in SPMS patients. These processes appear analogous to those observed in RRMS,.[16] suggesting that “plexus-related” inflammation may persist throughout the disease course and is not limited to early or relapsing phases.[33]

In summary, while lesion burden is the primary determinant of brain atrophy, particularly in SPMS, CP-related chronic inflammation likely contributes to ongoing neurodegeneration across phenotypes. These findings support the view that brain damage from early lesion activity plays a key role in shaping disease trajectory, while smouldering, CP-linked inflammation continues to influence progression within each clinical stage.

Our study has several limitations. Although we classified MS phenotypes as distinct groups, it is widely recognised that RRMS and SPMS are part of a clinical continuum with blurred boundaries.[34] However, our cohorts differ clearly in age and disease duration, supporting their distinction. Another limitation is that while NBV is a useful marker of brain atrophy,[35] it is a single time-point measure and does not reflect the dynamic nature of ongoing atrophy. Furthermore, cross-sectional analyses cannot fully capture the slow, CP-related progression that contrasts sharply with the acute, lesion-driven brain tissue damage occurring earlier in the disease course, potentially rendering the effect of CP on atrophy in SPMS less apparent. Additionally, confounders such as comorbidities and treatment effects were not fully accounted for and may influence CP volume and brain atrophy. Future longitudinal studies will be essential to better define CP-mediated mechanisms in MS.

## Conclusion

This study reveals three key insights into CP volume in MS. First, CP enlargement progresses across disease stages from CIS to SPMS. Second, it occurs independently of ventricular expansion, indicating distinct underlying pathological processes. Third, in SPMS, lesion burden drives damage, but CP-related inflammation continues to contribute to disease progression through its association with chronic lesion expansion and ongoing brain atrophy.

These findings position the CP as a potential biomarker and therapeutic target. Future longitudinal studies, particularly in SPMS, are needed to clarify its role in disease progression.

## Supporting information

All Supp Data

## Data Availability

All data produced in the present study are available upon reasonable request to the authors

## Declaration of conflicting interest

All authors declare no conflict of interest.

## Funding statement

Study was supported by Multiple Sclerosis Australia grant 214010.

## Data Availability Statement

Anonymised data is available upon reasonable request.

---

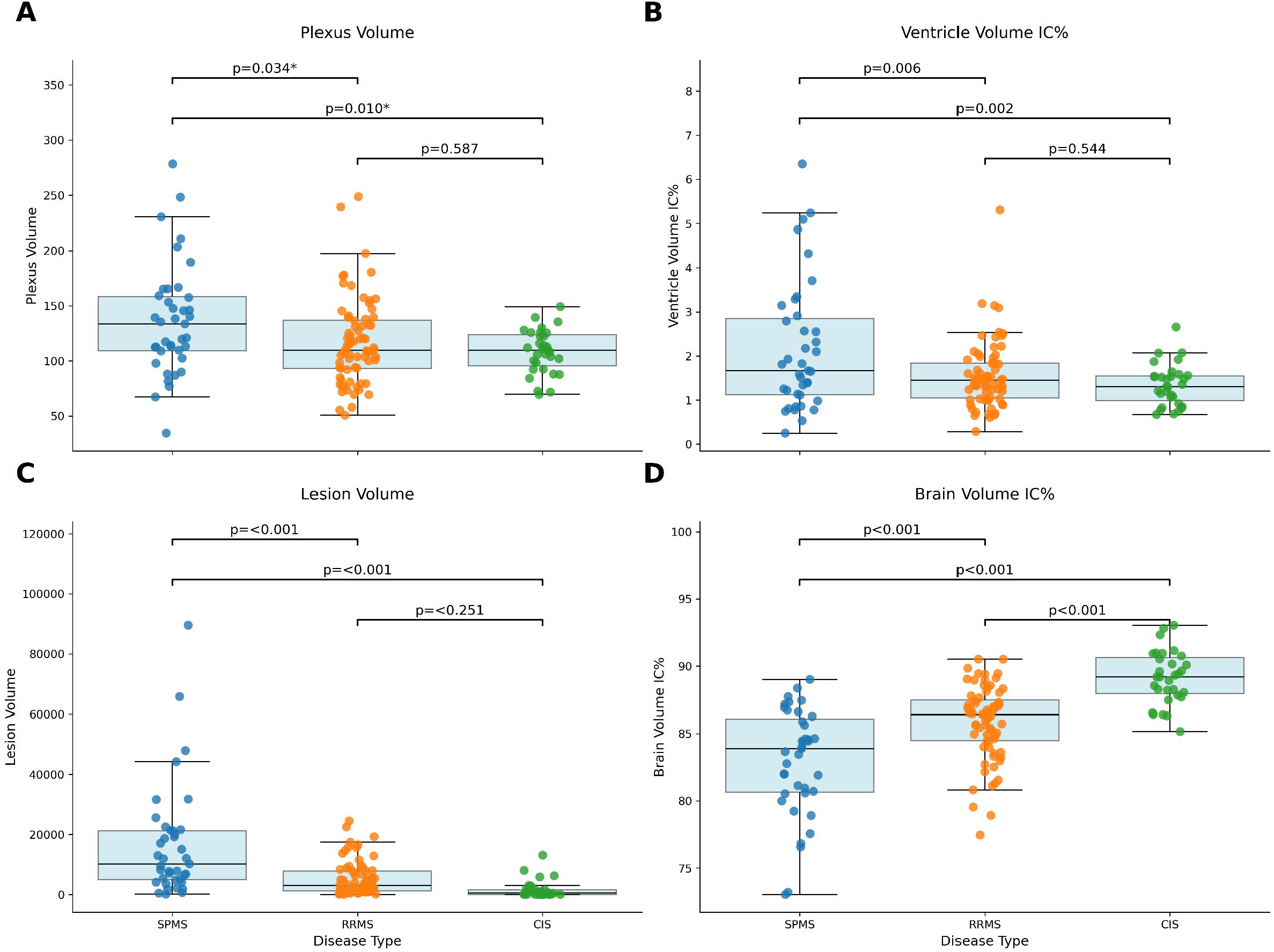

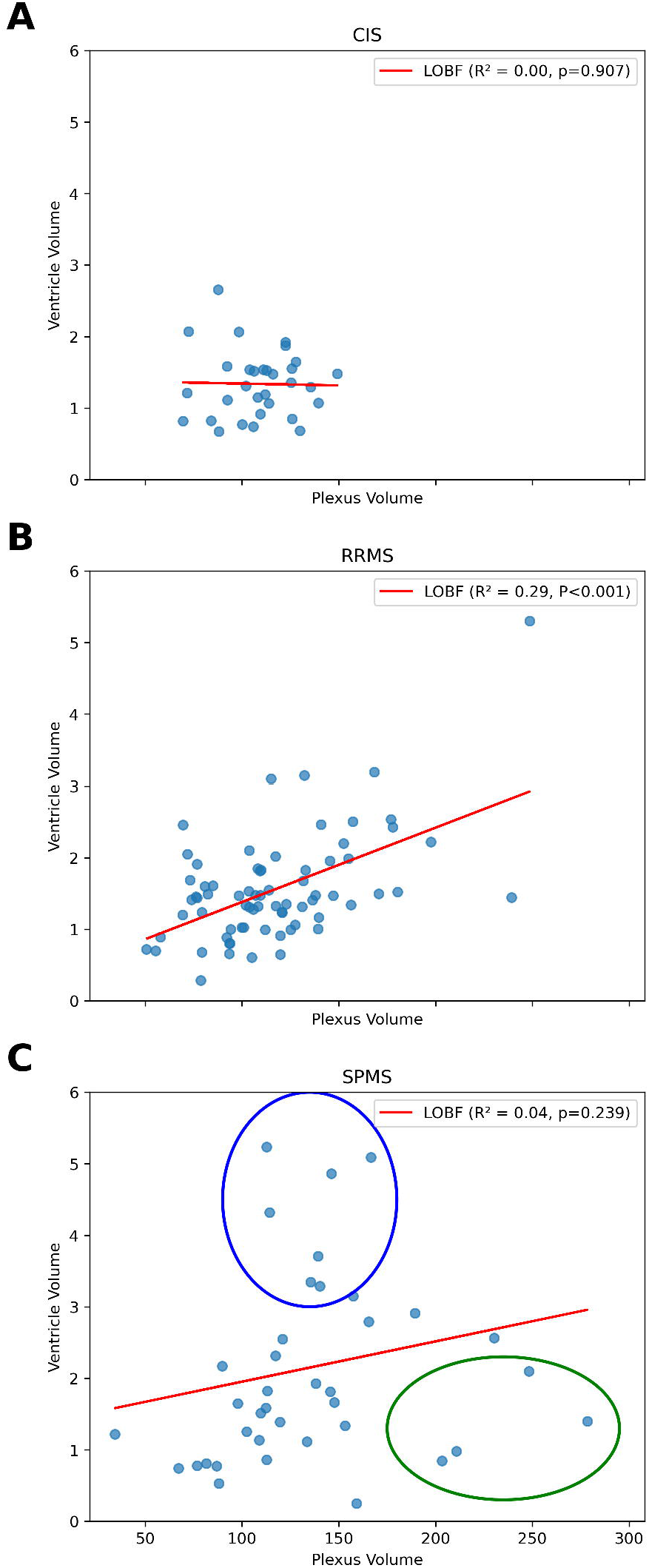

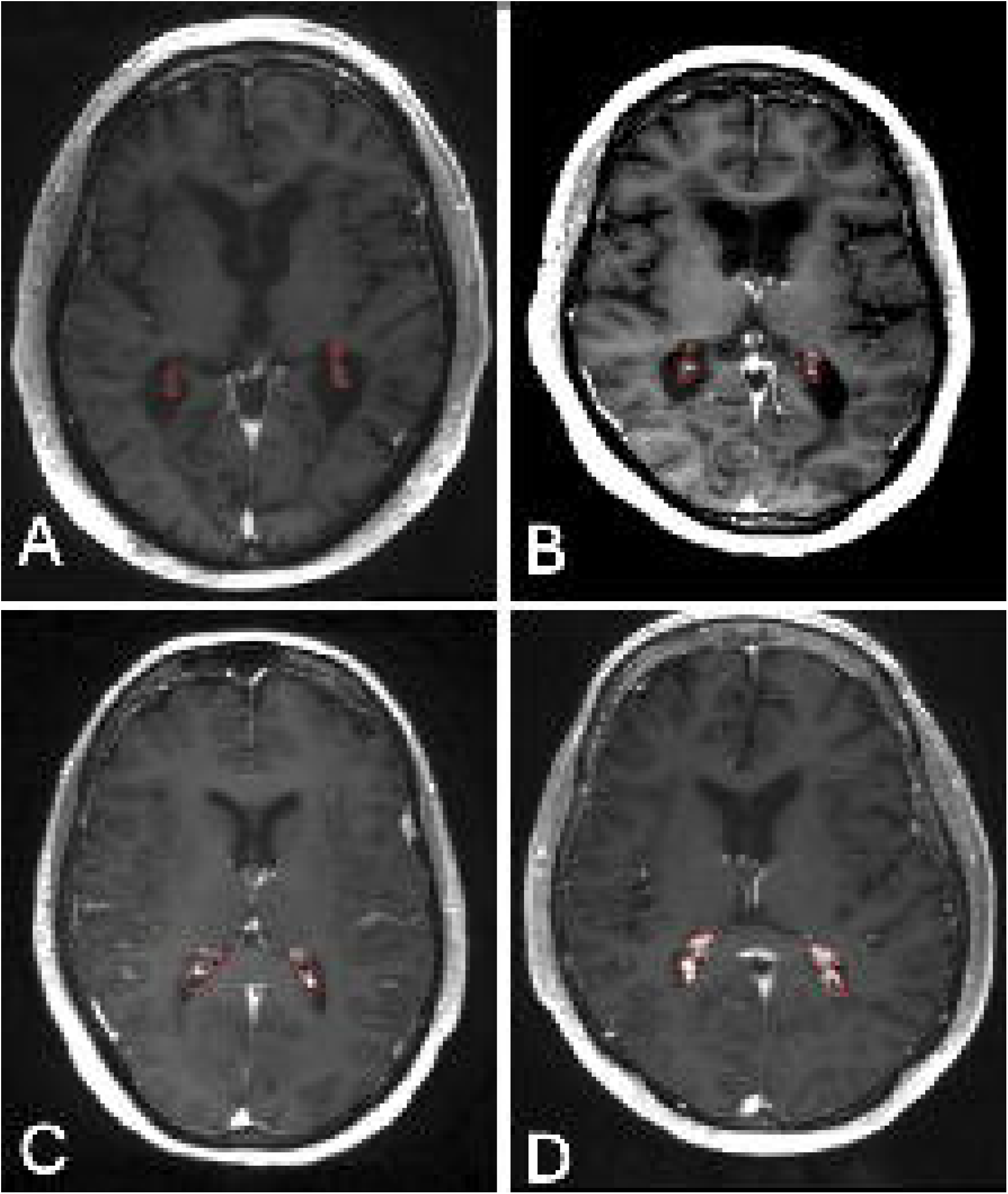

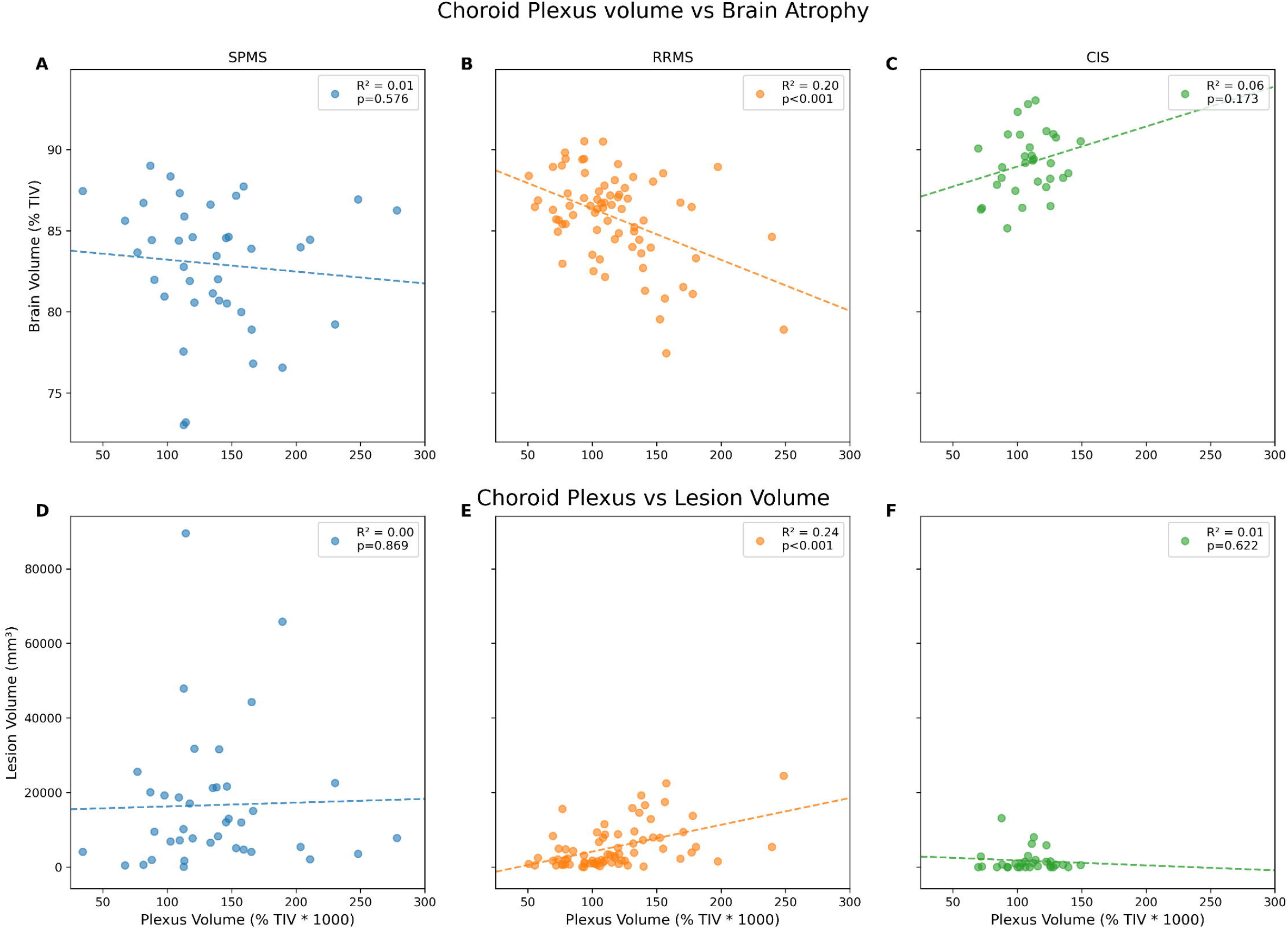

## Notes

### Competing Interest Statement

The authors have declared no competing interest.

### Funding Statement

This work was funded my MS Australia

### Author Declarations

This study was approved by University of Sydney Human Research Ethics Committees and followed the tenets of the Declaration of Helsinki. Written informed consent was obtained from all participants.

