## Supplementary material for "Choroid Plexus Enlargement in Secondary Progressive MS: phenotype comparison": All Supp Data

**Specific MRI parameters and image processing.**

The following MRI sequences were acquired:

a. Pre- and post-contrast (gadolinium) Sagittal 3D T1: GE BRAVO sequence, duration 4 min each, FOV 256mm, Slice thickness 1mm, TE 2.7ms, TR 7.2ms, Flip angle 12°, Pixel spacing 1mm. The acquisition Matrix (Freq x Phase) is 256x256, which results in 1mm isotropic acquisition voxel size. The reconstruction matrix is 256x256.
b. FLAIR CUBE; GE CUBE T2 FLAIR sequence, duration 6 min, FOV 240mm, Slice thickness 1.2 mm, Acquisition Matrix (Freq x Phase) 256x244, TE 163ms, TR 8000ms, Flip angle 90°, Pixel spacing 0.47 mm. The reconstruction matrix is 512x512.
c. Echo-Planar Imaging based diffusion-weighted MRI, duration 9 min (64-directions with 2 mm isotropic acquisition matrix, TR/TE = 8325/86 ms, b = 1000 s/mm^2^, number of b0 = 2).

**MRI image pre-processing:**

The baseline T1-weighted imaging was realigned to Anterior and Posterior Commissure (AC-PC) orientation. Using FLIRT (FSL, FMRIB Software Library), follow-up T1 images were co-registered to initial (month 0) AC-PC space by applying transformation matrices derived from linear co-registration between baseline AC-PC aligned brain and follow-up native T1 brain images. In parallel, diffusion MRI was corrected for motion and eddy-current distortion in FSL, and then EPI susceptibility distortion was minimised by applying deformation maps generated from nonlinear co-registration between DWI b0 brain images and T1-weighted images at each time-point using ANTs (Advanced Normalization Tools). Subsequently, tensor reconstruction was performed in MRtrix3. Tensor and FLAIR images were then linearly co-registered to corresponding T1 AC-PC images at each time point.

**Age-related adjustments**

Age-related effects were addressed through distinct adjustment procedures derived from healthy control cohorts. For choroid plexus (CP) volumes, we first characterised the relationship between age and CP volume in normal controls using quadratic regression analysis (as shown in Fig 2 [1]). This yielded:

$$CP_{LOBF}= 0.021\bullet age^{2}- 0.64\bullet age + 74.06$$

Where:

- $CP_{LOBF}: reference CP volume of a healthy subject \left( \% of TIV\bullet1000 \right)$
- $age: subjects age in years$

Analysis revealed a minimum at 15.6 years (offset 61.9), incorporated into age adjustment as:

$$CP_{Adjusted}= CP_{measured}- CP_{age}-CP_{offset}$$

Where:

- $CP_{adjusted}: age-adjusted CP volume (\% of tiv\bullet1000)$
- $CP_{measured}: subject's measured CP volume (\% of TIV\bullet1000$
- $CP_{age}: calculated CP volume derived from the quadratic function$
- $CP_{offset} : minimum point (61.9) at age 15.6$

For ventricular volumes, an analogous approach yielded:

$${VV}_{LOBF}= 0.00094\bullet age^{2}- 0.062\bullet age + 2.22$$

Where:

- ${VV}_{LOBF}: reference ventricle volume of a healthy subject \left( \% of TIV \right)$

Analysis revealed a minimum at age 33 (offset 1.19), incorporated as:

$${VV}_{Adjusted}= {VV}_{measured}- {VV}_{age}-{VV}_{offset}$$

Where:

- ${VV}_{adjusted}: age adjusted ventricle volume (\% of tiv)$
- ${VV}_{measured}: subject's measured ventricle volume (\% of TIV)$
- ${VV}_{age}-represents the calculated ventricle volume derived from the quadratic function$
- ${VV}_{offset}: minimum point (1.19) at age 33$

Brain volume adjustment followed methods from Study B [reference]. For subjects over 35 years:

$${Brain}_{LOBF}= 0.00176\bullet age+93.85$$

Where:

- ${Brain}_{LOBF}: reference brain volume of a healthy subject (\% of tiv)$

Brain age adjustment was then calculated as:

$${Brain}_{Adjusted}= {Brain}_{measured}- {Brain}_{age}-{Brain}_{offset}$$

Where:

- ${Brain}_{Adjusted}: age adjusted brain volume (\% of TIV)$
- ${Brain}_{measured}: subject's measured brain volume (\% of TIV)$
- ${Brain}_{age}: calculated brain volume derived from the linear function$
- ${Brain}_{offset}: reference point \left( 87.7 \right) at age 35$

**Figures**

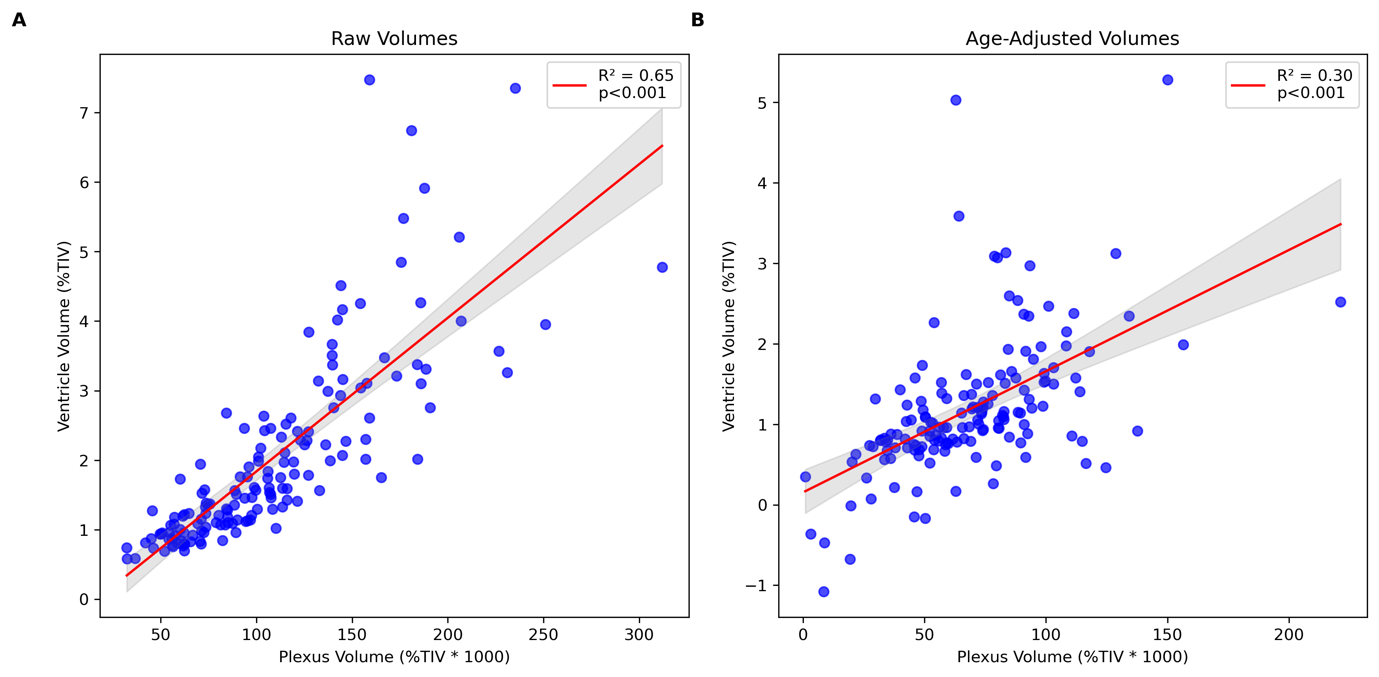

Supplementary Figure 1

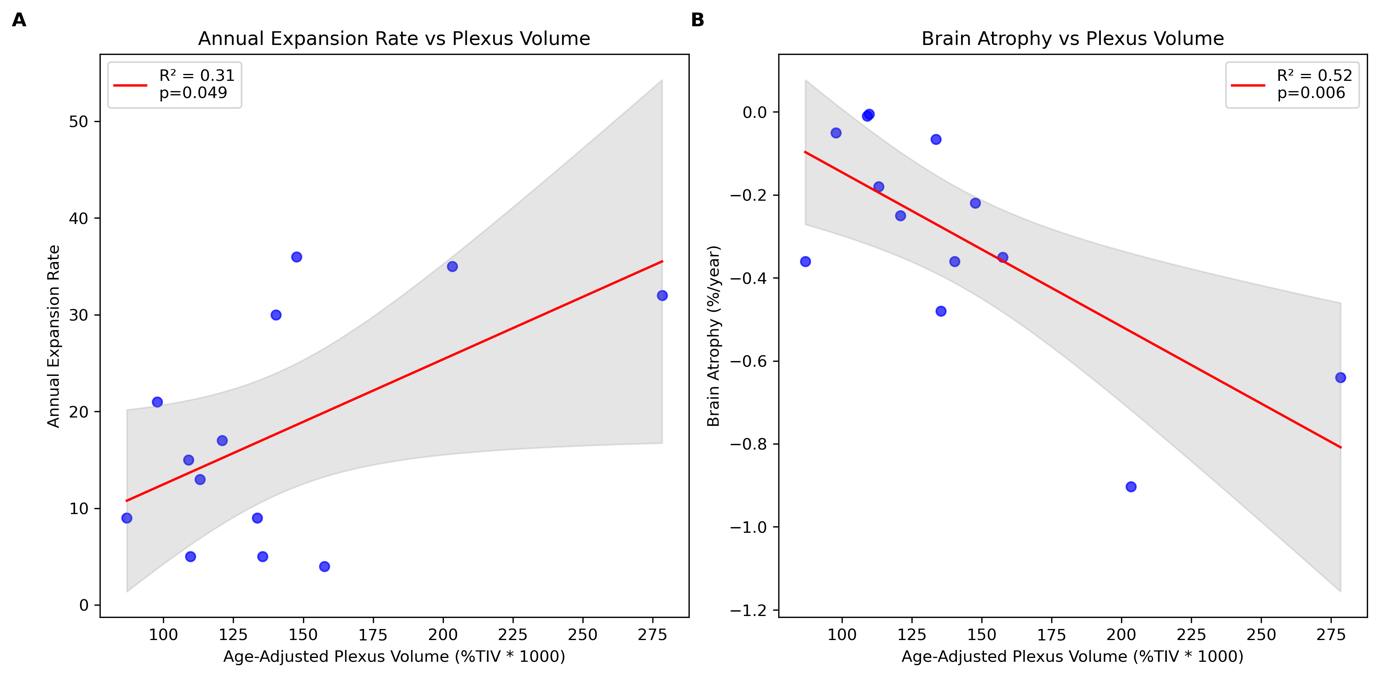

Supplementary Figure 2
